# HOW COMMUNITY PHARMACIES CAN RAISE UTERINE CANCER AWARENESS AMONG WOMEN FROM ETHNIC MINORITY POPULATIONS

**DOI:** 10.64898/2026.03.23.26349058

**Authors:** H Omar, A Chitrakar, M Patel, N Darko, EL Moss

**Affiliations:** College of Life Sciences, University of Leicester, Leicester, UK, LE2 7LX; University Hospitals of Leicester NHS Trust, Infirmary Square, Leicester, LE1 5WW; Centre for Research Equity, University of Oxford, Oxford, OX2 6GG

**Keywords:** uterine cancer, red-flag, symptoms, community pharmacies, ethnic minority

## Abstract

**Background:** Community pharmacies are a potential route to reach underserved groups due to their locations and staff, who often reflect the diversity of the populations they serve. This study aimed to explore the potential for a uterine cancer awareness campaign encouraging earlier presentation of red-flag symptoms amongst ethnic minority women conducted through community pharmacies.

**Methods:** Semi-structured qualitative interviews were conducted with pharmacy staff. Views and experiences on the role of the pharmacy in cancer awareness, the impact of language and cultural sensitives on discussions, and signposting for red-flag symptoms were explored using a pre-designed interview schedule. Interviews were analysed using reflexive thematic analysis.

**Results:** Fifteen pharmacy staff (pharmacists, technicians, and managers) working in a variety of clinical settings participated in an interview. The identified themes were: diversity and demographics; the community and its pharmacy; and women’s health. All participants reported low awareness of uterine cancer and its red-flag symptoms, however, saw the potential for pharmacies to reach women in their communities. A number of participants reported prior experience in sign-posting symptomatic patients to primary care but lacked confidence to discuss red-flag symptoms due to their limited knowledge. Pharmacists identified the need for formal standardised training for staff to ensure accuracy knowledge dissemination and mobilisation.

**Conclusion:** This study has shown the huge potential of community pharmacies to address cancer inequities through culturally tailored, co-designed interventions. However, structured staff education, ensuring accuracy of information disseminated, and formal signposting or pathways for symptomatic patients enabling timely onward referral for investigation would be needed.

## INTRODUCTION

Significant disparities have been identified in the stage of presentation of uterine cancer amongst women from Black ethnic groups [1], which consequently results in worse long-term survival [2]. Previous research has identified barriers to accessing healthcare including a lack of awareness of cancer symptoms [3] and the suspected cancer referral processes [4]. A lack of awareness of uterine cancer, of which endometrial cancer accounts for approximately 95% of cases, and the normalisation of red flag symptoms are also commonly reported [5]. Symptoms are often misattribution to benign conditions, in particular uterine fibroids, or false reassurance from cervical screening [6]. Discussion of gynaecological conditions is often viewed as very personal or even taboo in some communities, with women reluctant to openly discuss symptoms or concerns, and this can add to a lack of confidence or embarrassment in seeking medical review [7]. The misconception that uterine cancer is synonymous with cervical cancer and can arise as a result of sexual behaviour can compound stigmas, resulting in further isolation and delayed presentation. In addition, a personal view of low perceived risk or a fatalistic view towards cancer can also delay presentation of symptoms [8]. Multi-lingual, culturally competent information materials have been shown to be value assets in changing health seeking behaviours [9].

The role and reach of community pharmacies has changed considerably over the past two decades [10]. Community pharmacies have long been identified as a potential route to reach underserved populations due to their locations, with 98.3% of the English urban population having a community pharmacy within a 20-minute walk [11], however erosion of the ‘positive pharmacy care law’ has been reported due to a reduction in the community pharmacy availability, especially in locations of greater socioeconomic deprivation [12]. To date the role for community pharmacies to support cancer awareness and early cancer diagnosis has been limited, however, Public Health England in 2021 outlined a roadmap in which opportunities might be utilised to address health inequalities which included the use of local pharmacies due to their locations within communities [13]. This has been further expanded in the 10 year NHS plan, which strongly emphasises prevention, early diagnosis and an expanded role for community pharmacies [14].

A uterine cancer awareness campaign aimed at women from Black and Asian minority populations involving outreach events in North East London identified that the reach within target communities was limited [15]. Consequently, a short animated uterine cancer awareness video called ‘Seeing red..?’ was co-designed with women from ethnic minority groups, uterine cancer survivors and healthcare professionals including information on red-flag symptoms, next steps, risk factors and addressing common misconceptions [16]. The versatility of the ‘Seeing red..?’ raised the potential for uterine cancer awareness dissemination through community pharmacies, particularly those located within areas with high ethnic minority populations.

A study was conducted to explore the opinions of staff working within community pharmacists, technicians, and sales staff to understand their views and experiences of the potential challenges and barriers to a uterine cancer awareness intervention utilising the ‘Seeing red..?’ multilingual information materials delivered through community pharmacies.

## METHODS

Ethical approval was granted by the University of Leicester Research Ethics Committee (reference number 39250). Recruitment for the study was through professional networks and snowball recruitment. Individuals interested in participating in the study were sent an invitation letter and the participant information sheet information via email and had the opportunity to ask questions of the study team. All participants provided written informed consent before undertaking a one-to-one virtual interview (Microsoft Teams) with a member of the research team.

A structured interview schedule was followed designed in collaboration with a leader in pharmacy practice and policy development. Firstly, the participants’ background and experience of cancer awareness and women’s health was explored, particularly amongst women from an ethnic minority group. Secondly, the participant was shown the ‘Seeing red..?’ animated uterine cancer awareness video, and feedback requested. Lastly, questions were asked as to how such information resources could be disseminated through community pharmacies, along with potential facilitators and barriers. It was acknowledged that the term ‘ethnic minority’ was an umbrella term used to describe a very diverse populations and that there is great heterogeneity within and between groups.

The interviews were led by two female researcher students who were trained in qualitative techniques, HO a medical student undertaking a MSc, (supported by ELM) and AC, a specialist gynaecology doctor undertaking a PhD. The researchers had no prior relationship with the participants however, the study invitation letter introduced the two researchers and that the study was part of their research degrees. The interviews were transcribed using Microsoft Teams and field notes were taken.

Data were analysed using a reflexive thematic analysis, as described by Braun and Clarke [17]. There were no repeat interviews, transcripts were not returned to the participants for checking and did not provide feedback on the findings. Three key aspects to be included in the analysis were identified *a priori* from pilot work: pharmacist experience, uterine cancer awareness, and barriers to displaying or disseminating information. New themes and sub-themes were identified from the data, and added to pre-identified themes, in order to develop the finalised themes.

### Reflexivity and researcher positionality

Reflexivity was embedded throughout the study in line with guidance for reflexive thematic analysis (Braun and Clarke, 2022) and broader methodological guidance on reflexive practice in research [18]. Interviews were conducted by female researchers with clinical training backgrounds, which may have influenced data collection through shared professional language or assumptions regarding cancer awareness and care pathways. To mitigate this, interviewers adopted an open, non-directive approach, encouraged participants to elaborate on their perspectives, and avoided affirming clinical assumptions during interviews.

During analysis, reflexive discussions were used to challenge interpretations, particularly where early coding reflected clinical expectations rather than participants’ accounts. Codes and themes were iteratively reviewed within the wider multidisciplinary research team, which included researchers with expertise in social science, health equity, and inclusive research practice. This broader perspective supported critical reflection on assumptions and helped ensure that interpretations remained grounded in participants’ accounts rather than professional expectations.

Initial coding was undertaken by the interviewers (HO/AC) independently, who were closely familiar with the data through conducting the interviews and transcription. Coding was conducted inductively, with codes developed to capture patterned meaning across the dataset rather than to achieve inter-coder reliability. Codes and candidate themes were then iteratively discussed within the wider multidisciplinary research team, enabling refinement, re-naming, and re-structuring of themes through reflexive dialogue.

Theme development was an ongoing, recursive process, with themes reviewed in relation to the coded data and the full dataset to ensure coherence and analytic depth. Qualitative data management was supported using NVivo, which facilitated organisation of transcripts and codes but did not replace interpretive analysis by the research team. This approach is consistent with reflexive thematic analysis, where rigour is demonstrated through transparency and reflexive engagement rather than replication or coder agreement (Braun and Clarke, 2022).

## RESULTS

### Participants

In total, 15 individuals, representing 15 separate pharmacies from different locations across the UK, who were invited agreed to an interview between February 2024 and March 2025. A number of participants self-identified as belonging to an ethnic minority group and/or speaking a language in addition to English. The total number of people who received an invitation to participate was not known due to the open call through professional networks. The median interview duration was 30 minutes, ranging from 30 to 50 minutes. Of the 15 participants, the majority worked in a community pharmacy (60%) and were female (80%) (Table 1).

**Table 1.** Demographic characteristics and pharmacy location of study participants. Co-located - indicates works in a community pharmacy co-located with a GP practice.

| Participant | Gender | Role in Healthcare |
| --- | --- | --- |
| P1 | M | Community pharmacist (senior manager) |
| P2 | F | Community and hospital pharmacist |
| P3 | M | Community and hospital pharmacist |
| P4 | F | Hospital pharmacist |
| P5 | F | Hospital pharmacist |
| P6 | F | Community pharmacy technician (trainee) |
| P7 | F | Community pharmacy technician |
| P8 | F | Community pharmacist (retired) |
| P9 | F | Community pharmacist |
| P10 | M | Community pharmacist |
| P11 | F | Community pharmacist |
| P12 | F | Community pharmacist |
| P13 | F | Community pharmacist (co-located) |
| P14 | F | Community pharmacist (co-located) |
| P15 | F | Community pharmacist (co-located) |

Thematic analysis led to the identification of three main themes: Diversity and Demographics; The Community and its Pharmacy; and Women’s Health (Table 2). The themes identified through analysis of the interviews acknowledged the pivotal role that community pharmacies and pharmacists play, especially within ethnic minority communities, and identified the key requirements that would be required for a future pharmacy-based awareness campaign, and have been visually represented in Figure 1 and Figure 2.

**Figure 1.**
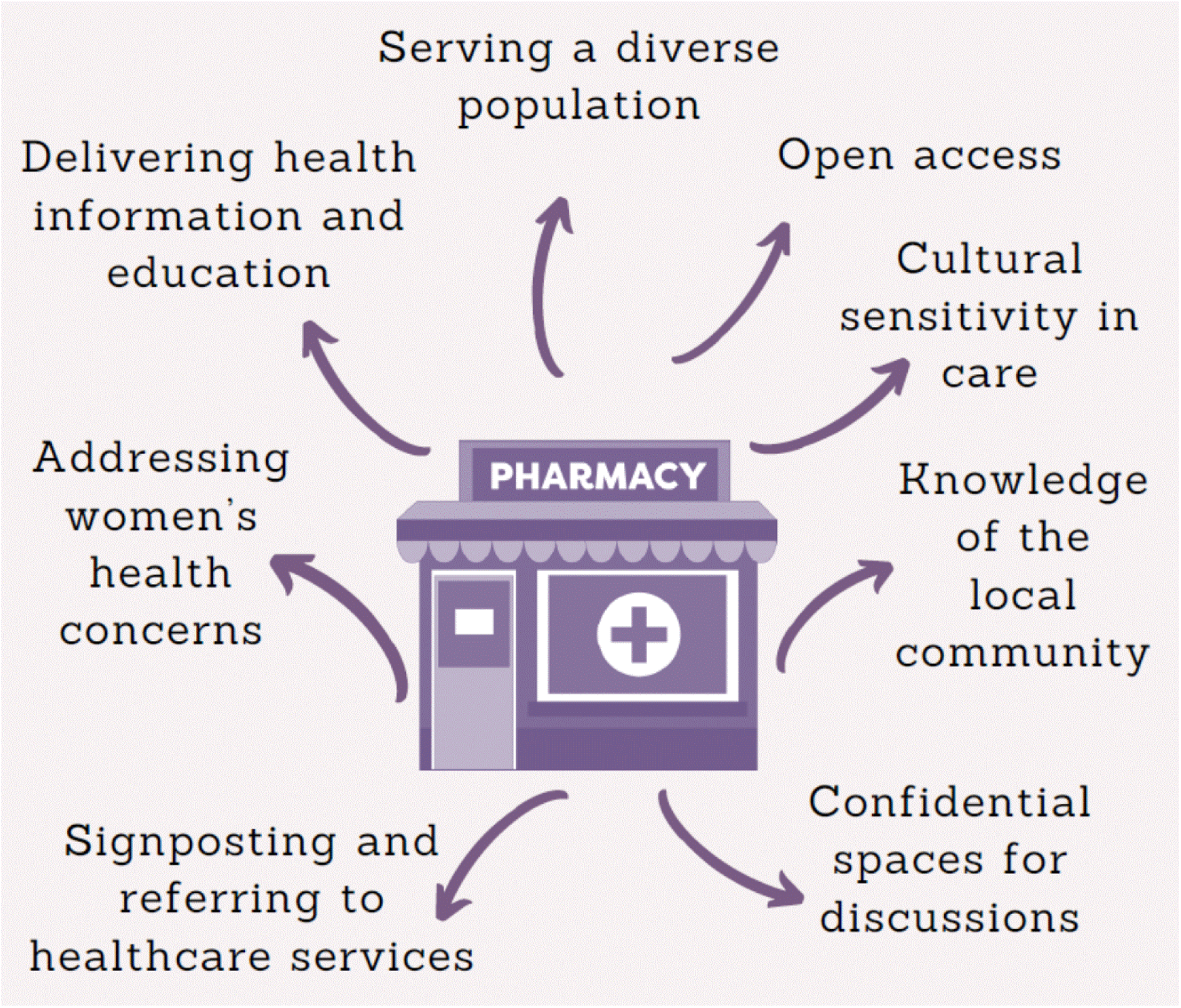
The many and diverse ways that community pharmacies can support women’s health and uterine cancer awareness.

**Figure 2.**
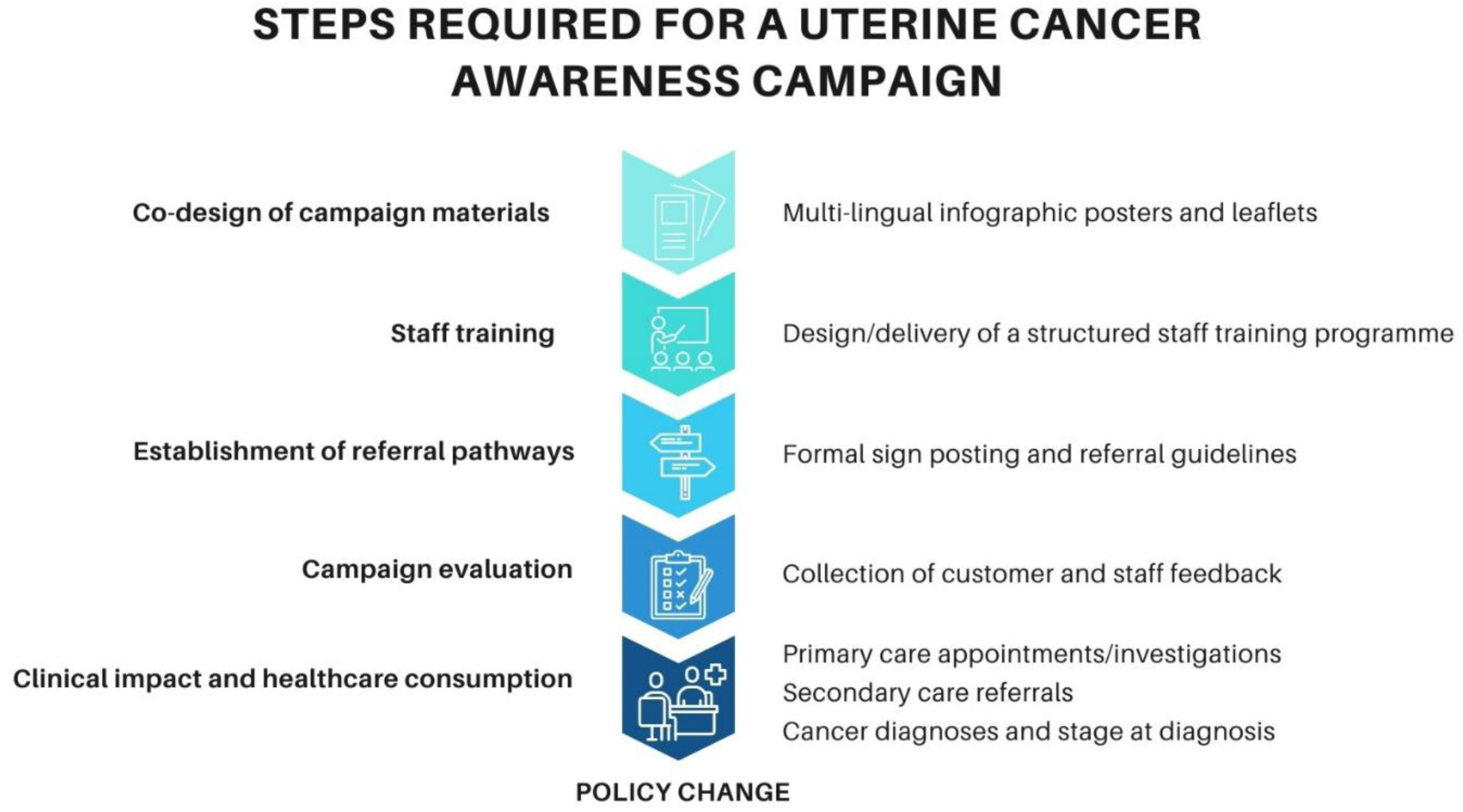
Steps required for a uterine cancer awareness campaign delivered through community pharmacies.

**Table 2:** Summary of themes and subthemes.

| **Themes** | **Sub-themes** |
| --- | --- |
| Diversity and Demographics | Diversity in service users |
|  | Challenges that can arise |
|  | Overcoming obstacles |
| The Community and Its Pharmacy | Role of community pharmacy staff |
|  | Relationship with service users |
|  | Accessibility of services and campaigns |
| Women's Health | Lack of awareness |
|  | Lack of discussion |
|  | Importance of safe space |

### Diversity and Demographics

This theme refers to the differences that participants observed amongst the service users that frequent community pharmacies, the potential challenges for delivery of care to a population with diverse needs and the efforts made by community pharmacy staff to accommodate for these differences (Table 2). The ethnic diversity of people attending for community pharmacy services was confirmed and participants agreed that people attending a community pharmacy reflected the local population: ‘*Because the pharmacy is based on Asian area, most of our patients are Asian’* (P6).

The diversity amongst service users was identified to bring challenges in access and delivery and of healthcare, with language in particular, being a potential barrier to imparting information, causing concerns amongst pharmacists when trying to communicate risks or side effects of medications: *‘[In] some areas it’s really difficult because not many people know English’* (P9)

However, many participants reported that community pharmacy staff often spoke the same languages as their service users, negating the need for translation services. Cultural beliefs and practices held by ethnic minority groups were also viewed as influential potentially affecting access to care at community pharmacies and ‘*more than [the] language barrier’* (P5). The need for cultural competency was acknowledged, in some cases with staff giving patients additional information in order to address cultural views that contradicted medical information. This included a preference for women to speak to female staff members about health concerns, and the understanding amongst pharmacists that a lack of availability could result in a lost consultation opportunity. Therefore, although barriers were identified that influenced the access of service users, participants described ways in which community pharmacy staff work overcome these barriers to deliver the care needed.

These findings align with UK conceptualisations of cultural competence, in which responsiveness to individuals’ cultural values, language needs, and expectations is central to effective engagement within community healthcare settings [18]. Participants’ accounts suggest that shared language and cultural familiarity enhance acceptability of services, positioning community pharmacies as culturally responsive access points for diverse underserved populations.

### The community and its pharmacy

This theme focused on how community pharmacies impact the wider community and the resources they provide for their service users, developing a unique bond with the local community and a point of access to the services and campaigns.

The interviews revealed the numerous and varied tasks and roles undertaken by pharmacists and within a community pharmacy, far beyond merely dispensing medications. The unrestricted access to pharmacies without the need for an appointment for many and varied health concerns was raised by many participants, and raising awareness and signposting for medical conditions was identified as a prominent aspect of a community pharmacy’s work. The need for staff to recognise red flag symptoms and give factually accurate information was viewed as important, since it was acknowledged that this could lead to earlier diagnosis and improved prognosis for patients, who may have delayed contacting healthcare services due to a lack of awareness of symptoms.

Participants shared that community pharmacy staff worked closely with the local population, forming a relationship with the wider community that differed from other healthcare providers, especially GP practices, with more frequent visits often leading to the development of a rapport between the staff and the local lay population. The relationship between pharmacy staff and the local community was felt to be strengthened because many staff came from the communities that they served, reflecting local recruitment strategies.

The wide variety of services offered by community pharmacies and the lessons learned from health awareness campaigns to date, for example blood pressure checks and Pharmacy First, were reported to be evidence supporting a potential role for pharmacies to act as hubs for sharing cancer awareness information. It was felt that even passive dissemination of information through displayed materials would prompt the public to engage in conversations and request more details, as seen with Pharmacy First, without requiring staff to actively disseminate information or adding to their workload.

From a health behaviour perspective, community pharmacies provide an important *opportunity* for engagement by offering an easily accessible, familiar environment that lowers structural and psychological barriers to help-seeking [19]. However, participants’ emphasis on passive dissemination highlights a tension between this opportunity and the *capability and motivation* of staff to initiate proactive health discussions, indicating that the feasibility of awareness activities is closely tied to their integration within existing pharmacy workflows.

### Women’s Health

This theme encompassed community pharmacy staff’s understanding and attitudes towards uterine cancer, the approach to women’s health within community pharmacies, and how pharmacies can be utilised to accommodate and facilitate conversations that may arise as a result of displaying uterine cancer information materials.

Participants in general reported a lack of knowledge of uterine cancer and its red-flag symptoms, with some even admitting that they had *‘zero knowledge on uterine cancer’* (P10). There was also the common misconception that cervical screening was also able to detect uterine cancer, and several individuals were of the view that regular smear tests might protect women from developing uterine cancer.

The participants reported that the ‘Seeing red..?’ video increased their understanding of uterine cancer, highlighting the video’s utility in raising awareness amongst both lay and clinical staff. Participants felt that there was an expectation of pharmacy staff to answer questions on a wide range of issues, however, it was acknowledged that they could not be expected to have in-depth knowledge of every medical condition, and knowledge was reported to typically reflect the priorities of service users. Therefore, short education videos, such as ‘Seeing red..?’, as part of a continuing professional development programme could be used to address knowledge deficits and increase the information accuracy and the confidence of staff to share with the public seeking information. Participants did highlight the need to improve women’s healthcare in general, including menopause care and cervical screening uptake, through greater training opportunities for pharmacists, thereby empowering staff with the knowledge to play an active role in improving the patient care.

Many participants held the view that women’s health was not commonly or widely discussed. This was attributed not only to women’s health being given lower priority compared to men’s health issues, but also to cultural and traditional expectations that discouraged open discussion of health concerns. In particular, gynaecological issues were reported to be a sensitive subject, with women were reluctant to discuss concerns or symptoms and it was felt that when women eventually spoke about female-related health concerns it was because symptoms were difficult to self-manage and were having an impact on their quality of life. Therefore, whilst women do talk to community pharmacy staff about gynaecological conditions, these conversations tend to be reactive, due to symptoms, rather than preventative. Embarrassment was cited as a common obstacle preventing women from engaging with pharmacy staff but that this could be overcome by creating a welcoming environment and initiating discussions asking about their symptoms. Uterine cancer red-flag symptoms, in particular postmenopausal bleeding, was viewed as ‘sensitive’ but community pharmacy staff were competent in navigating private conversations, facilitated by the use of a pharmacy consultation room. Participants’ reported lack of confidence to discuss uterine cancer reflects limitations in perceived *capability*, rather than unwillingness to engage. Within a COM-B framework, this suggests that while community pharmacies offer the *opportunity* for preventive conversations, insufficient training constrains staff *motivation* to initiate discussions, reinforcing the need for structured education to support proactive and accurate information sharing [19].

## DISCUSSION

The results of this study support the potential of community pharmacies as knowledge hubs for cancer awareness information, in particular in reaching individuals from underserved or non-English speaking communities [20] and open new avenues for pharmacy services as part of health service reforms. Community pharmacies have been identified as accessible health care locations that have enhanced ‘cultural hub’ potential for reaching ethnically diverse local population, facilitating health promotion programs tailored to community need [21]. Displaying uterine cancer awareness information and signposting to clinical services was well received by the study participants, supporting previous work that identified with adequate training and resources, pharmacy professionals can readily engage in cancer awareness programs, thereby increasing public awareness and screening uptake [22].

Interpreted through the COM-B model of health behaviour, the findings suggest that community pharmacies create strong *opportunity* for engagement through accessibility, familiarity, and trust, particularly for underserved populations [19]. However, limitations in staff *capability* (knowledge and confidence) and competing demands on *motivation* arising from workload pressures constrain proactive cancer awareness activity. These findings indicate that effective implementation will depend on interventions that strengthen staff capability through training and align awareness activities with existing commissioned services to ensure sustainability.

The increasing a burden and significantly worse uterine cancer survival amongst women from ethnic minority groups is a growing public health concern in the UK [23]. Previous cancer awareness campaigns have utilised mass media, such as TV and radio, and celebrity endorsements, however, these have been shown to have a lower impact amongst ethnic minority and non-English speaking populations [24], indicating the need for culturally-tailored interventions [25], as demonstrated with the ‘You need to know’ campaign’ [15]. Community pharmacies, being easily accessible and viewed as reliable, are in a good position to effectively support such interventions [21]. Previous research on pharmacy-led cancer awareness has been encouraging, for example, the ‘Be Clear on Cancer’ bowel screening awareness campaign, piloted in pharmacies, reported increased public awareness and greater screening uptake among target groups, especially when pharmacy staff were trained and supported with appropriate materials [26].

Our study has also highlighted community pharmacies’ role in health promotion and addressing misconceptions, especially amongst ethnic minority groups since staff often have language skills and cultural competencies that align with the local population [27]. Pharmacy staff have been recognised for their ability to provide culturally competent care, which is essential in effectively communicating health information to diverse populations [28]. Language is a considerable barrier to both accessing and receiving healthcare [29], however this appears to be less of a challenge within community pharmacies due to staff’s multilingual abilities, which may not be present in other healthcare sectors [30]. Many study participants reported fluency in non-English languages commonly spoken within the communities they serve, including Gujarati, Punjabi, Swahili and Arabic, thereby reducing the need for translation services, and facilitating patient understanding and adherence to medical advice [31]. The highest uterine cancer mortality rates have been reported in women of Black ethnicity and therefore ensuring that the language preferences of this population are included in awareness materials is essential. Additionally, awareness of cultural practices or ‘taboos’, separate from addressing language needs, reportedly enables the avoidance of misunderstandings [32], and helps to build trust with the service users [33]. This cultural competence is crucial in engaging in uterine cancer awareness discussions and can leverage the skills that pharmacy staff have developed through experience in advising on other medical conditions [34].

The sharing of uterine cancer awareness does not propose any extension of diagnostic or treatment responsibilities within community pharmacy. The activity described is limited to public health awareness, recognition of potential red-flag symptoms, and signposting to appropriate healthcare services, all of which are established components of community pharmacy practice [35]. Professional boundaries for an awareness intervention would be maintained through the absence of diagnostic interpretation or clinical decision-making, and through clear communication that pharmacy advice does not replace medical assessment. From a medico-legal perspective, these activities are already governed by existing professional standards, duty of care, and indemnity arrangements, and therefore do not introduce new legal or professional risk. Any potential risk of misinformation arises from unstructured discussions rather than the pharmacy setting itself and is mitigated through the use of standardised, evidence-based information, staff training, and clear escalation pathways. Recognition of red-flag symptoms and onward signposting are embedded within existing pharmacy governance frameworks and align with national clinical guidance [36], functioning solely as an early awareness and access mechanism that complements, rather than replaces, medical assessment pathways. Formal training for staff on the risk factors and signs/symptoms of uterine cancer would be essential for a future campaign, since the self-reported knowledge of study participants was low [37]. One of the challenges in achieving early diagnosis is a lack of awareness of the red-flag symptoms [4], normalisation of symptoms, and a lack of confidence or embarrassment to seek medical review [6]. Co-created information materials within community spaces and on social media can increase awareness, as was demonstrated with the ‘You need to Know’ campaign [15]. Infographics are a commonly used information dissemination format used by pharmacies and supported by NHS England to share information [13] and could be utilised as part of a uterine cancer awareness campaign. Additionally, the barriers reported by patients in accessing primary care could potentially be overcome by the use of community pharmacies, due to consultations not requiring to be prescheduled, thereby enabling impromptu discussions [11]. Referral or signposted pathways would also need to be established, enabling prompt onward referral of individuals with red-flag symptoms. Previous cancer campaigns within pharmacies were successful when pharmacists received adequate training and referral guidelines appropriate to their local practice [22].

The role of the pharmacist in the UK has expanded considerably over the past two decades with increased NHS commissioning of extended roles, most notably with Pharmacy First [38]. Internationally many healthcare systems do not use this role, however since the aim of our study was to explore the potential of knowledge dissemination through pharmacies, rather than pharmacists undertaking an extended clinical role in triaging or diagnosing patients with red-flag symptoms, the results could be of interest outside the UK. The potential increase in consultations would have to be considered however, since many pharmacists did report that their workload had increased with the adoption of extended services, in particular under schemes such as Pharmacy First. Future implementation plans would need to factor in existing pressures and the time for awareness activities in a way that supports, rather than overwhelms, workload resulting in slower response time and truncated discussions [39]. Given that community pharmacists in the UK are already commissioned and reimbursed by the NHS/government to deliver certain public health programmes (e.g., diabetes prevention, smoking cessation, vaccination), similar approaches could be used to integrate women’s health awareness into established funding and service models.

The main limitation of this study was the sample size. Diverse backgrounds and geographical and working environments, aimed to maximise the range of opinions and experiences collected however, we do not have a breakdown of the ethnic groups served by the participants. In addition, recruitment continued until data saturation, and no new themes were generated. Since the study was primarily focusing on raising awareness amongst women from ethnic minority populations there was a risk of self-selection bias with participants that had a greater interest in this topic and/or self-identified as belonging to an ethnic minority group, which could potentially reduce the generalisability of the results. The study required participants to consider how a uterine awareness campaign could be implemented through community pharmacies and lacked real-world experiences, however the opinions of the pharmacists, although theoretical, have given valuable insights that can be used to inform the potential and design of a future campaign. The addition of patient and public perspectives on uterine cancer awareness through community pharmacies will be explored in future work and will focus on accessibility and utilisation.

## CONCLUSIONS

This study adds to the growing evidence that community pharmacies, through culturally informed and co-designed interventions, can contribute meaningfully to health equity and uterine cancer awareness. Structured training for pharmacy staff on red-flag symptoms and pathways for onward referral would need to be established, along with infographic resources to support passive dissemination. Further work is now needed to pilot such a campaign and investigate the potential impact on pharmacists’ workload as well as clinical utility and knowledge dissemination.

## Informed Consent Statement

All participants gave written informed consent before participating in the study.

## AI tools

No generative AI and AI-assisted technologies were used during the conduct of this study or in the manuscript preparation process.

## Institutional Review Board Statement

Ethical approval was granted by the University of Leicester Research Ethics Committee (reference number 39250). Date of approval 10/02/2023.

## Data Availability

The data underlying this article cannot be shared publicly due to the privacy of individuals that participated in the study. The data will be shared on reasonable request to the corresponding author.

## Acknowledgments

We authors wish to thank all the participants.

